# COVID-19 vaccine response in people with multiple sclerosis

**DOI:** 10.1101/2021.07.31.21261326

**Authors:** Emma C Tallantyre, Nicola Vickaryous, Valerie Anderson, Aliye Nazli Asardag, David Baker, Jonathan Bestwick, Kath Bramhall, Randy Chance, Nikos Evangelou, Katila George, Gavin Giovannoni, Leanne Grant, Katharine E Harding, Aimee Hibbert, Gillian Ingram, Meleri Jones, Angray S Kang, Samantha Loveless, Stuart J Moat, Neil P Robertson, Klaus Schmierer, Sita Navin Shah, Jessica Simmons, Matthew Upcott, Mark Willis, Stephen Jolles, Ruth Dobson

## Abstract

**Objective:** To investigate the effect of disease modifying therapies on serological response to SARS-CoV2 vaccines in people with multiple sclerosis

**Methods:** 473 people with multiple sclerosis from 5 centres provided one or more dried blood spot samples and questionnaires about COVID-19. Information about disease and drug history was extracted from their medical records. Dried blood spots were eluted and tested for antibodies to SARS-CoV2 receptor binding domain. Seropositivity was expressed according to validated cut-off indices. Antibody titers were partitioned into tertiles using data from people on no disease modifying therapy as a reference. We calculated the odds ratio of seroconversion (Univariate logistic regression) and compared quantitative vaccine response (Kruskal Wallis) following SARS-CoV2 vaccine according to disease modifying therapy. We used regression modelling to explore the effect of factors including vaccine timing, treatment duration, age, vaccine type and lymphocyte count on vaccine response.

**Results:** Compared to no disease modifying therapy, the use of anti-CD20 monoclonal antibodies (odds ratio 0.03; 95% confidence interval 0.01-0.06, p<0.001) and fingolimod (odds ratio 0.41; 95% confidence interval 0.01-0.12) were associated with lower seroconversion following SARS-CoV2 vaccine. All other drug groups did not differ significantly from the untreated cohort. Time since last anti-CD20 treatment and total time on treatment were significantly related with response to vaccination. Vaccine type significantly predicted seroconversion, but not in those on anti-CD20 medications.

**Interpretation:** Some disease modifying therapies carry a risk of attenuated response to SARS-CoV2 vaccination in people with MS. We provide recommendations for the practical management of this patient group.

## Background

Individuals with inflammatory neurological disorders have faced considerable uncertainty during the coronavirus disease 2019 (COVID-19) pandemic, related to both the potential risks posed by their underlying medical disorder, and the immunomodulatory treatment needed to manage their disease. Factors that appear to confer increased vulnerability for poor outcomes following symptomatic COVID-19 include advanced disability, obesity, male sex and the use of immunosuppressive drugs, in particular anti-CD20 monoclonal antibodies [1]. As a result, many people with chronic neurological disease were advised to strictly self-isolate during periods of high community transmission.

The development of effective vaccines against COVID-19 infection, which are safe for use in vulnerable groups, offer hope for a reduction in COVID-19 related mortality and morbidity. However, uncertainties remain over vaccine efficacy for those taking immunosuppressive and/or immunomodulatory drugs including multiple sclerosis (MS) disease modifying therapies (DMTs). Previous studies have demonstrated a blunted vaccine response in people with MS (pwMS) receiving ocrelizumab [2] and fingolimod [3]; however vaccine responses to a truly novel biological antigen (i.e. one giving rise to human disease, rather than keyhole limpet hemocyanin, KLH) have only been subject to limited study. Data is urgently required on the efficacy of COVID-19 vaccination for those individuals receiving immunomodulating drugs to guide them regarding risks related to reopening policies, and to inform policy around booster vaccinations [4] or additional strategies for those who are unable to respond adequately to vaccination. There may be the potential to optimise vaccination response in people receiving MS DMTs e.g. by temporarily suspending continuous DMT or extending the interval between intermittent DMTs. However, adopting this approach in the absence of supportive evidence needs to be balanced against unnecessary and potentially disabling MS disease activity.

Study of MS treatment offers a relatively unique opportunity. PwMS tend to remain on continuous platform (i.e. non-induction) DMTs for many years, rather than having treatment “holidays” or breaks, due to the irreversible nature of disease-related disability. In addition, DMTs are given as monotherapy, rather than in combination as is practice in some other autoimmune diseases and selected haematological malignancies. Furthermore, untreated pwMS are generally considered to have a “normal” immune response to vaccination. As a result, response to vaccination in pwMS offers an opportunity to understand the role of individual medications, and their mode(s) of action on the development of an adequate vaccine response.

In this study we investigate the effect of MS DMTs on the serological response to COVID-19 vaccination in a large multicentre cohort. Remotely patient-collected, posted dried blood spots were used to obtain samples from people with MS for antibody evaluation following vaccination. By establishing the antibody response to COVID-19 vaccines in relation to patient, treatment, and vaccine characteristics we aim to better inform future guidance for people with MS on vaccination policy, DMT management and infection protection.

## Methods

PwMS from five UK MS centres (Cardiff, Newport, Nottingham, Royal London Hospital (Barts Health NHS Trust) and Swansea) were invited to participate in this study. Samples were analysed in two laboratories: University Hospital of Wales (UHW), Cardiff and Queen Mary University of London (QMUL).

### Participants

Participants were asked to provide one or more dried blood spot samples along with a questionnaire, providing data on isolation behaviour, employment setting, clinical or laboratory confirmed cases of COVID-19 infection, vaccine dates and type. Participants were also asked to allow access to their medical records, allowing verification of MS diagnosis, details of DMT start dates, doses, timing and delay for vaccination. Hospital records were also used to provide confirmation of reported COVID-19 requiring hospital admission and to extract laboratory results such as total lymphocyte counts from routine blood monitoring data. All participants provided written informed consent to take part in this study.

This study has Research Ethics Committee approval (REC 19/WA/0058 (Wales REC 3 – covering samples processed in Cardiff), 05/WSE03/111 (South East Wales REC – covering samples processed in Cardiff) and 20/NE/0176 (Newcastle North Tyneside REC – covering QMUL samples)).

### Sample collection

Dried blood spot sampling was undertaken remotely, avoiding the need for phlebotomist time and hospital visits by potentially immunosuppressed patients. A kit containing instructions on how to collect samples, lancets to obtain capillary blood, a blood spot collection device (PerkinElmer 226 Spot Saver or Whatman 903 Protein Saver card) and a return mail envelope was posted to participants at their home address. Participants were asked to date their dried blood spot test card and return it using first class surface mail as soon as possible. Samples received at the UHW were stored desiccated at -20°C until analysis; those received at QMUL were stored at room temperature for a maximum of a week prior to spot punching and elution, with subsequent storage at -20°C.

### Antibody testing for SARS-CoV-2 in dried blood spot specimens

Dried blood spots were used to determine serological status in two laboratories; 376 in UHW and 97 in QMUL.

In UHW, samples were analysed with the COVID-SeroKlir two-step enzyme-linked immunosorbent assay (ELISA) (Kantaro Biosciences, USA - supplied by EKF Diagnostics, UK), which detects SARS-CoV-2 specific IgG antibodies against the receptor-binding domain (RBD) [5] [6], using resuspended dried blood spots, using previously validated methods [7]. A 6mm diameter sub-punch was taken from each dried blood spot using a DELFIA dried blood spot Puncher (PerkinElmer) and placed into 2ml 96 well plates (Waters #186002482) containing 600μl of the Kantaro Kit sample diluent (equivalent to 1:100 dilution recommended for the Kantaro kit). The 96 well plates were then covered and shaken at room temperature for 1 hour [as we have shown previously this to be the optimum time to ensure complete extraction of the blood from the filter paper collection device]. The plates were then stored at 4°C until analysis (usually within 72 hours), although extracted samples were stable for at least 1 week. One hundred microlitres of the dried blood spot eluate was transferred to the 96-well ELISA plate and analysed on an automated platform (DSX, Dynex Technologies, USA) according to the manufacturer’s instructions.

The Kantaro assay relies on an assay-specific calibrator to report the ratio of the specimen absorbance to the calibrator absorbance to calculate a cut-off index (CI) value. The antibody level in plasma / serum is reported as positive (≥0.7) or negative (<0.7). In a series of paired plasma and dried blood spots (n=22 antibody negative and n=25 antibody positive samples) we observed good agreement between paired specimens (r=0.994) and the gradient of the slope was 1.241x. Based upon the negative bias observed in dried blood spot vs paired plasma samples we calculated the following CI for dried blood spot specimens – positive (≥0.56) and negative (<0.56). Quality controls (QC) as supplied by the manufacturer and in-house prepared dried blood spot QC’s (positive and negative) were analysed on each assay plate.

At QMUL, samples were analysed using the Globody technique [8], with the RBD as the target antigen. A 4mm blood spot punch was eluted in 200ul of 1% Triton X-100 in PBS at room temperature overnight. Eluted samples were assayed the next day or stored at -20C until analysis. Assays were carried out using 20ul of dried blood spot eluate mixed with 30ul of a 50% slurry of Protein G agarose equilibrated in PBS (Thermofisher Scientific), made up to a final volume of 1.2ml with 0.05% PBS-Tween 20 (PBST) and left for 30min on a rotating wheel at room temperature. A PBST wash was performed by pelleting the agarose at 200xg for 1 minute, aspirating supernatant down to ∼100ul and washing with 1100ul PBST. After washing, an additional 200xg for 1 min spin was performed and aspiration of non-agarose bound mixture to ∼100ul. RBD-GloBody preparation (8.7ul)of 1×10^8^ lux units was added to each sample and then made up to 750ul with PBST and left on rotating wheel for 30min, after which the agarose resin was applied to a Pierce 0.8ml centrifuge column (Thermofisher Scientific). Columns were washed five times with 750ul PBST to remove unbound RBD-GloBody and followed by a final wash with 750ul PBS. Bound IgG and RBD-GloBody in complex with IgG were eluted from spin column with the addition of 100ul of 0.1M Glycine, pH 2.7 and neutralised with 12ul 0.1M Tris pH 9.0. Assays on 30ul of the resulting eluate were performed in triplicate using furimazine substrate (20ul furimazine in 1ml 0.1% BSA in PBS) (Promega). Luminescence was measured on a CLARIOstar plate reader. Validation work confirmed a cut-off index (CI) value for dried blood spots of positive (≥0.56) and negative (<0.56). Quality controls (QC) as supplied by the manufacturer and in-house prepared dried blood spot QCs (positive and negative) were analysed on each assay plate. A limit of blank (LoB) was determined by LoB=mean luminescence:blank + 2.58 (standard deviation:blank), values greater than this suggest seroconversion with a confidence of 99%.

### Statistical Analysis

Participants were categorised according to DMT exposure status at the time of first COVID-19 vaccination. People were considered exposed if they had received alemtuzumab or cladribine within 4 years, ocrelizumab within 12 months, natalizumab within 8 weeks, fingolimod, dimethyl fumarate, teriflunomide, glatiramer acetate or beta-interferon within 4 weeks of their vaccine first dose. All other participants were categorised as unexposed to DMT.

Fishers exact test was used to compare the chance of seroconversion following first and second vaccine dose across different DMTs. Univariate logistic regression with no DMT as the reference group was used across DMT groups to provide odds ratio of seroconversion by DMT class.

Quantitative vaccine responses were partitioned into tertiles with no DMT as the reference group. Kruskal Wallis test was used to compare the proportion of vaccine responses in each tertile between DMT following vaccine 2.

The impact of time between last dose of anti-CD20 monoclonal antibody (ocrelizumab and rituximab, excluding ofatumamab given different administration schedule) and first vaccine dose, and odds of seroconversion, was established using univariate logistic regression. Similar analyses were performed in order to establish the impact of time since treatment initiation on odds of seroconversion. Linear regression across tertiles with the same outputs was performed in order to explore the impact on quantitative vaccine response.

Finally, stepwise multivariate logistic regression was used to model the likelihood of a positive antibody response to COVID-19 vaccine measured after the second vaccine dose. Two groups of interest were selected for study - (1) those either not on DMT, or taking DMT shown not to influence seroconversion in the univariate analyses, and (2) those who had been treated with anti-CD20 monoclonal antibodies. An initial model including gender and vaccine type was generated; subsequently age, EDSS (grouped into 0.0-4.0, 4.5-5.5, 6.0-6.5 and 7.0-10.0), previous COVID-19 symptoms, time between vaccine doses, time from second vaccine to sampling, total time on most recent DMT and lymphocyte count were added to the model in a stepwise manner. Only those factors shown to significantly improve model fit were retained in the final model. Mann-Whitney U test was used to compare potential influence on anti-SARSCoV2 IgG titre according to vaccine type.

Statistical analysis was performed in Stata v16 (Stat corp ltd).

## Results

473 participants returned at least one dried blood spot sample collection device and were included in the analysis. 58 participants provided samples prior to vaccination, 246 during the interval between their first and second vaccine dose, and 430 following the second vaccine dose. Demographic and clinical features are given in Table 1.

**Table 1:**
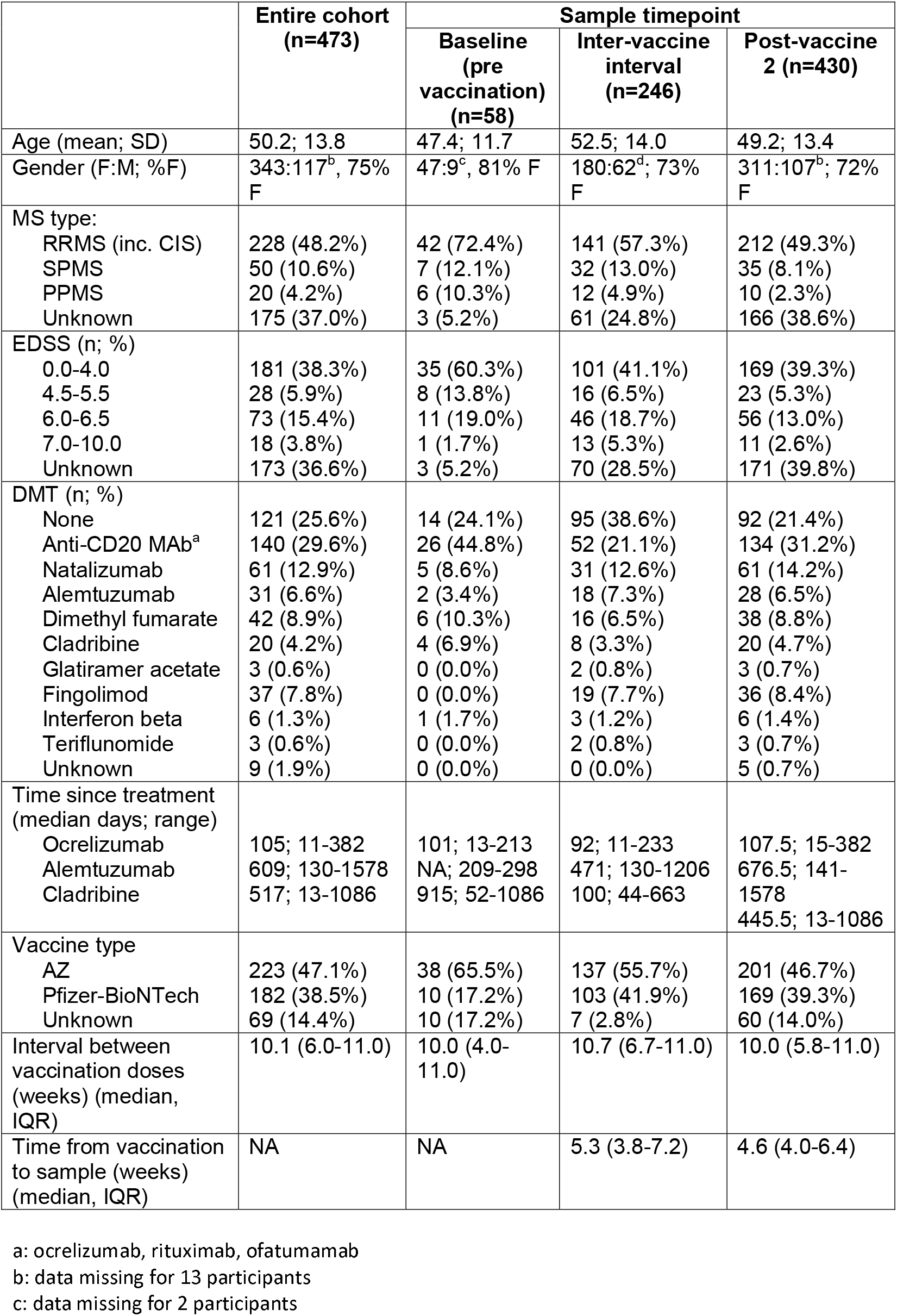
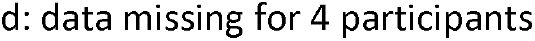
Clinical and demographic features of the study population

|  | Entire cohort<br>(n=473) | Sample timepoint |  |  |
| --- | --- | --- | --- | --- |
|  |  | Baseline<br>(pre<br>vaccination)<br>(n=58) | Inter-vaccine<br>interval<br>(n=246) | Post-vaccine<br>2 (n=430) |
| Age (mean; SD) | 50.2; 13.8 | 47.4; 11.7 | 52.5; 14.0 | 49.2; 13.4 |
| Gender (F:M; %F) | 343:117<sup>b</sup>, 75%<br>F | 47:9<sup>c</sup>, 81% F | 180:62<sup>d</sup>; 73%<br>F | 311:107<sup>b</sup>; 72%<br>F |
| MS type: |  |  |  |  |
| RRMS (inc. CIS) | 228 (48.2%) | 42 (72.4%) | 141 (57.3%) | 212 (49.3%) |
| SPMS | 50 (10.6%) | 7 (12.1%) | 32 (13.0%) | 35 (8.1%) |
| PPMS | 20 (4.2%) | 6 (10.3%) | 12 (4.9%) | 10 (2.3%) |
| Unknown | 175 (37.0%) | 3 (5.2%) | 61 (24.8%) | 166 (38.6%) |
| EDSS (n; %) |  |  |  |  |
| 0.0-4.0 | 181 (38.3%) | 35 (60.3%) | 101 (41.1%) | 169 (39.3%) |
| 4.5-5.5 | 28 (5.9%) | 8 (13.8%) | 16 (6.5%) | 23 (5.3%) |
| 6.0-6.5 | 73 (15.4%) | 11 (19.0%) | 46 (18.7%) | 56 (13.0%) |
| 7.0-10.0 | 18 (3.8%) | 1 (1.7%) | 13 (5.3%) | 11 (2.6%) |
| Unknown | 173 (36.6%) | 3 (5.2%) | 70 (28.5%) | 171 (39.8%) |
| DMT (n; %) |  |  |  |  |
| None | 121 (25.6%) | 14 (24.1%) | 95 (38.6%) | 92 (21.4%) |
| Anti-CD20 MAb<sup>a</sup> | 140 (29.6%) | 26 (44.8%) | 52 (21.1%) | 134 (31.2%) |
| Natalizumab | 61 (12.9%) | 5 (8.6%) | 31 (12.6%) | 61 (14.2%) |
| Alemtuzumab | 31 (6.6%) | 2 (3.4%) | 18 (7.3%) | 28 (6.5%) |
| Dimethyl fumarate | 42 (8.9%) | 6 (10.3%) | 16 (6.5%) | 38 (8.8%) |
| Cladribine | 20 (4.2%) | 4 (6.9%) | 8 (3.3%) | 20 (4.7%) |
| Glatiramer acetate | 3 (0.6%) | 0 (0.0%) | 2 (0.8%) | 3 (0.7%) |
| Fingolimod | 37 (7.8%) | 0 (0.0%) | 19 (7.7%) | 36 (8.4%) |
| Interferon beta | 6 (1.3%) | 1 (1.7%) | 3 (1.2%) | 6 (1.4%) |
| Teriflunomide | 3 (0.6%) | 0 (0.0%) | 2 (0.8%) | 3 (0.7%) |
| Unknown | 9 (1.9%) | 0 (0.0%) | 0 (0.0%) | 5 (0.7%) |
| Time since treatment<br>(median days; range) |  |  |  |  |
| Ocrelizumab | 105; 11-382 | 101; 13-213 | 92; 11-233 | 107.5; 15-382 |
| Alemtuzumab | 609; 130-1578 | NA; 209-298 | 471; 130-1206 | 676.5; 141-1578 |
| Cladribine | 517; 13-1086 | 915; 52-1086 | 100; 44-663 | 445.5; 13-1086 |
| Vaccine type |  |  |  |  |
| AZ | 223 (47.1%) | 38 (65.5%) | 137 (55.7%) | 201 (46.7%) |
| Pfizer-BioNTech | 182 (38.5%) | 10 (17.2%) | 103 (41.9%) | 169 (39.3%) |
| Unknown | 69 (14.4%) | 10 (17.2%) | 7 (2.8%) | 60 (14.0%) |
| Interval between<br>vaccination doses<br>(weeks) (median,<br>IQR) | 10.1 (6.0-11.0) | 10.0 (4.0-11.0) | 10.7 (6.7-11.0) | 10.0 (5.8-11.0) |
| Time from vaccination<br>to sample (weeks)<br>(median, IQR) | NA | NA | 5.3 (3.8-7.2) | 4.6 (4.0-6.4) |

a: ocrelizumab, rituximab, ofatumumab
b: data missing for 13 participants
c: data missing for 2 participants
d: data missing for 4 participants

Participants received their initial vaccine dose between 8 December 2020 and 9 June 2021 Vaccine type was available in 404 (85.4%) participants; 181 reported vaccination with an mRNA vaccine (167 BNT162b2 mRNA (Pfizer-BioNTech), 1 Johnson and Johnson) and 223 reported vaccination with the ChAdOx1 nCoV-19 (Oxford–AstraZeneca) adenoviral vector vaccine. One participant reported initial vaccination with the ChAdOx1 nCoV-19 (Oxford–AstraZeneca) adenoviral vector vaccine with the second vaccine dose BNT162b2 mRNA (Pfizer-BioNTech); this participant was excluded from analyses using vaccine type as a variable. Median interval between vaccine doses was 10.0 weeks (range 3.0-14.0 weeks; interquartile range (IQR) 6.1-11.0 weeks).

### Pre-vaccine serology

Of the 58 vaccine naïve baseline samples, 6 (10.3%) were seropositive and 52 (89.7%) seronegative for SARS-CoV2. Of the 6 participants who were seropositive at baseline; 4 were on DMT (1 glatiramer acetate, 1 alemtuzumab (year 1 only; April 2019) and 2 ocrelizumab). Data on prior symptoms suggestive of COVID-19 was available for 44 out of 58 participants, including all 6 who were seropositive at baseline. Only 3 out of 6 seropositive participants reported prior symptoms or prior positive SARSCoV-2 PCR. Eleven (19.2%) of the 52 individuals who were seronegative at baseline self-reported prior symptoms suggestive of COVID-19 (of whom 3 had confirmatory SARSCoV-2 PCR). There was no significant association between patient-reported historical COVID-19 infection and baseline seropositivity.

### Post-vaccine serology

Overall, 140/246 (56.9%) participants with a sample available between first and second vaccine dose demonstrated a positive IgG response (Table 2), and 280/430 (65.1%) at > 4 weeks post second dose. Univariate logistic regression demonstrated that, compared to no DMT, the use of anti-CD20 monoclonal antibodies (odds ratio (OR) 0.03; 95% confidence interval (95%CI) 0.01-0.06, p<0.001) and fingolimod (OR 0.41; 95%CI 0.01-0.12) were associated with lower seroconversion following vaccine 2 (Table 2). All other DMT did not differ significantly from the untreated cohort. Subgroup analysis between the UHW and QMUL analysed cohorts did not demonstrate any significant difference between the response to complete vaccination course in any DMT that passed the multiple testing threshold (Supplementary table 1).

**Table 2:** Vaccine response according to DMT

|  | Serostatus following dose 1 (positive:negative, % seroconverted) | Serostatus following dose 2 (positive:negative, % seroconverted) | OR seroconversion following dose 2 (OR; 95% CI) |
| --- | --- | --- | --- |
| All participants | 140:106 (57%) | 280:150 (65%) | NA |
| No DMT | 77:18 (81%) | 85:7 (92%) | 1 (reference) |
| Anti-CD20 MAb<sup>a</sup> | 7:45 (13%) | 33:101 (25%) | 0.03 (0.01-0.06)* |
| Natalizumab | 23:8 (74%) | 56:5 (92%) | 0.92 (0.28-3.05) |
| Alemtuzumab | 14:4 (78%) | 24:4 (86%) | 0.49 (0.13-1.83) |
| Dimethyl fumarate | 9:7 (56%) | 35:3 (92%) | 0.96 (0.23-3.93) |
| Cladribine | 1:7 (13%) | 16:4 (80%) | 0.33 (0.09-1.26) |
| Glatiramer acetate | 2:0 (100%) | 3:0 (100%) | NA<sup>b</sup> |
| Fingolimod | 4:15 (21%) | 12:24 (33%) | 0.04 (0.01-0.12)* |
| Interferon Beta | 3:0 (100%) | 5:1 (83%) | 0.41 (0.04-4.03) |
| Teriflunomide | 0:2 (0%) | 3:0 (100%) | NA<sup>b</sup> |
|  | **p<0.0001** | **p<0.0001** |  |

\* p<0.0001
a: ocrelizumab, rituximab, ofatumumab
b: logistic regression limited by small numbers

One-way ANOVA to examine the effect of DMT on antibody titre was limited by ceiling effects in the UHW cohort and small numbers in treatment groups when restricting to the QMUL cohort. Using tertiles defined by antibody titres in the untreated cohort, Kruskal-Wallis equality of populations rank test demonstrated a significant difference between populations in IgG titre tertile between DMT (Figure 1). This persisted regardless of whether both seropositive and seronegative samples were included in the analysis (Figure 1a, p=0.0001), or if the analysis was restricted to only the seropositive cohort (Figure 1b, p=0.0022).

**Figure 1:**
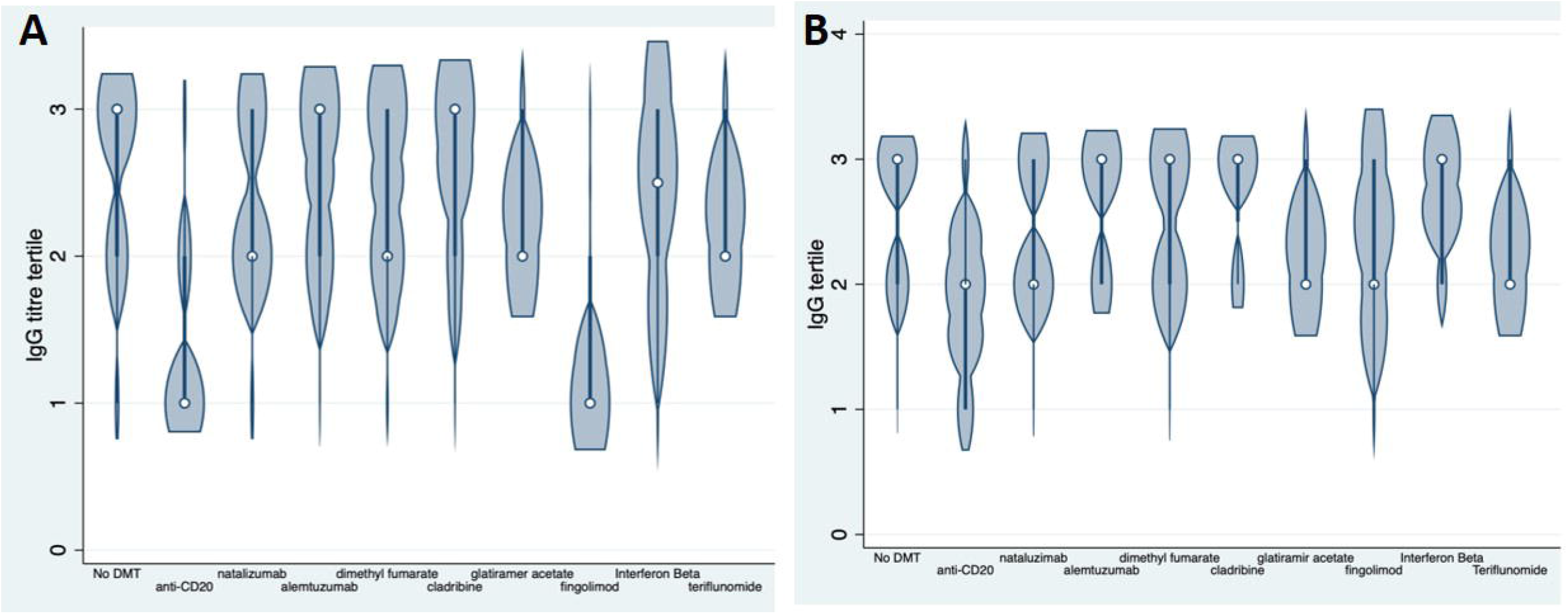
Quantitative vaccine response following complete vaccine course by DMT (a) All samples (b) Seropositive samples only

Neither time since last anti-CD20 treatment nor total time on anti-CD20 DMT predicted (binary) serostatus after second vaccine, however the small number of seropositive individuals limited the power in this analysis. 5/72 people treated with anti-CD20 therapies within the past 5 months mounted a response following the second vaccine dose, compared to 5/9 people treated >=7 months ago (p=0.037, Fishers exact test). Linear regression across tertiles, excluding a single outlier, was performed in the anti-CD20 cohort. In this analysis, both time from last treatment administration to initial vaccination (Figure 2a, p=0.033) and total time on treatment (Figure 2b, p=0.025) demonstrated a significant relationship with serological response to vaccination.

**Figure 2:**
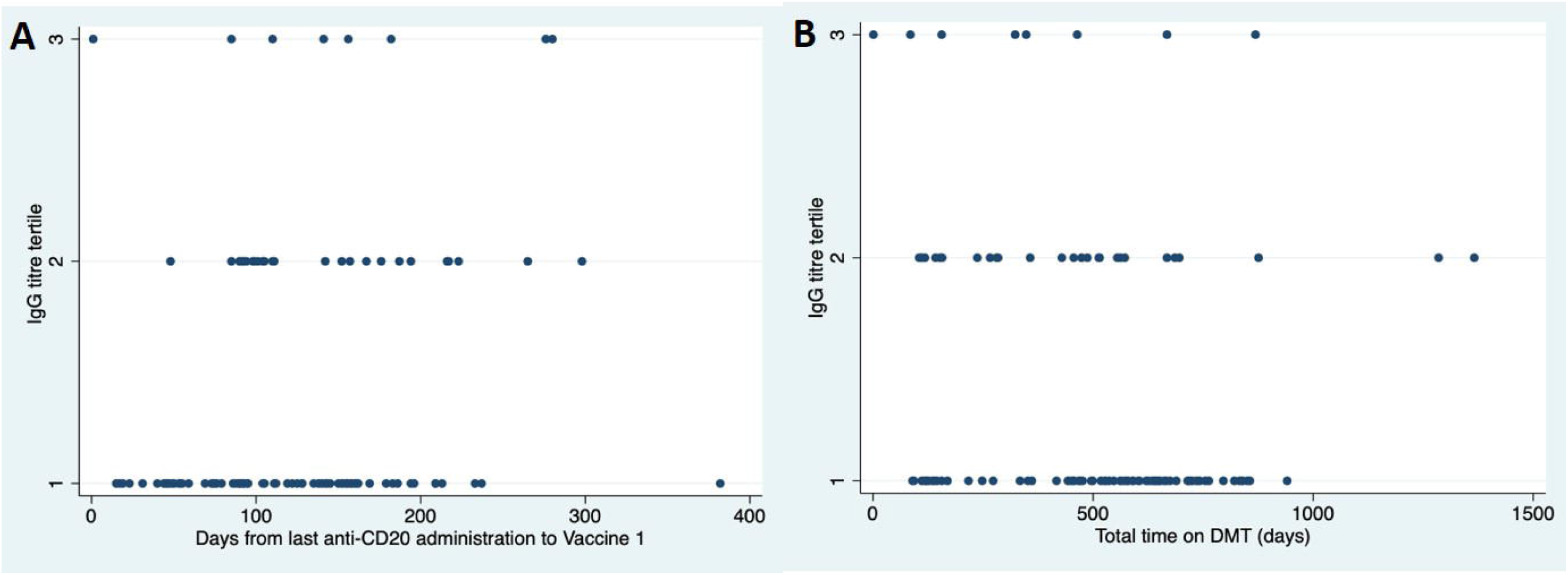
Scatter plot demonstrating relationship between (a) time from last treatment administration to first vaccine dose, and (b) total time on treatment and tertile of IgG response to vaccination

DMT type was then dichotomised into anti-CD20 monoclonal antibody (ocrelizumab, rituximab and ofatumamab) and control MS with normal vaccine response (no DMT, natalizumab, alemtuzumab, dimethyl fumarate, cladribine, glatiramer acetate, interferon beta, and teriflunomide; these DMT all demonstrated a statistically similar serological response to vaccination. Samples from fingolimod treated individuals were excluded from this analysis. In order to develop a multivariate regression model, a stepwise multivariate regression model was developed. Factors retained in the final, best performing model were vaccine type and treatment (anti-CD20 vs others). Whilst time between administration of second vaccine dose and age showed significance on initial testing, they did not pass the threshold for multiple testing on Bonferroni correction (p=0.005) (Table 3). Overall IgG levels were not included in this analysis as only 4/473 participants had levels below the lower limit of normal. Finally, the impact of vaccine type on SARS-CoV2-IgG titre was examined across DMT groups (anti-CD20 vs others). There was no difference between serological response to vaccination according to vaccine type in those who had received anti-CD20 monoclonal antibodies (Figure 3a, p=0.39). However, in the control group, those who were vaccinated with the BNT162b2 mRNA (Pfizer-BioNTech) vaccine had a significantly greater IgG response to vaccination than those vaccination with the ChAdOx1 nCoV-19 (Oxford–AstraZeneca) vaccine (Figure 3b, p<0.0001).

**Table 3:**
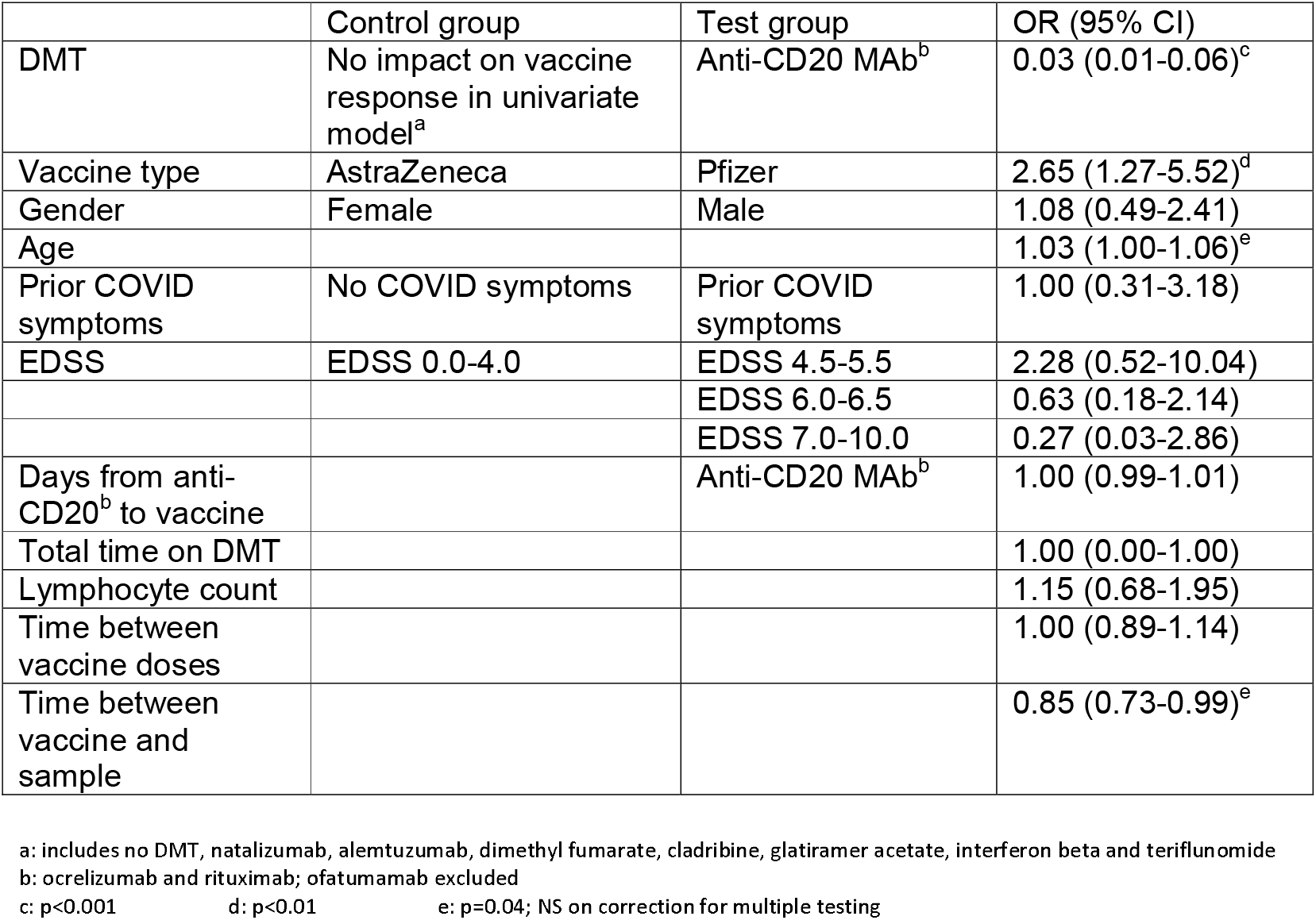
Factors predicting serostatus following vaccine dose 2 in best performing model

**Figure 3:**
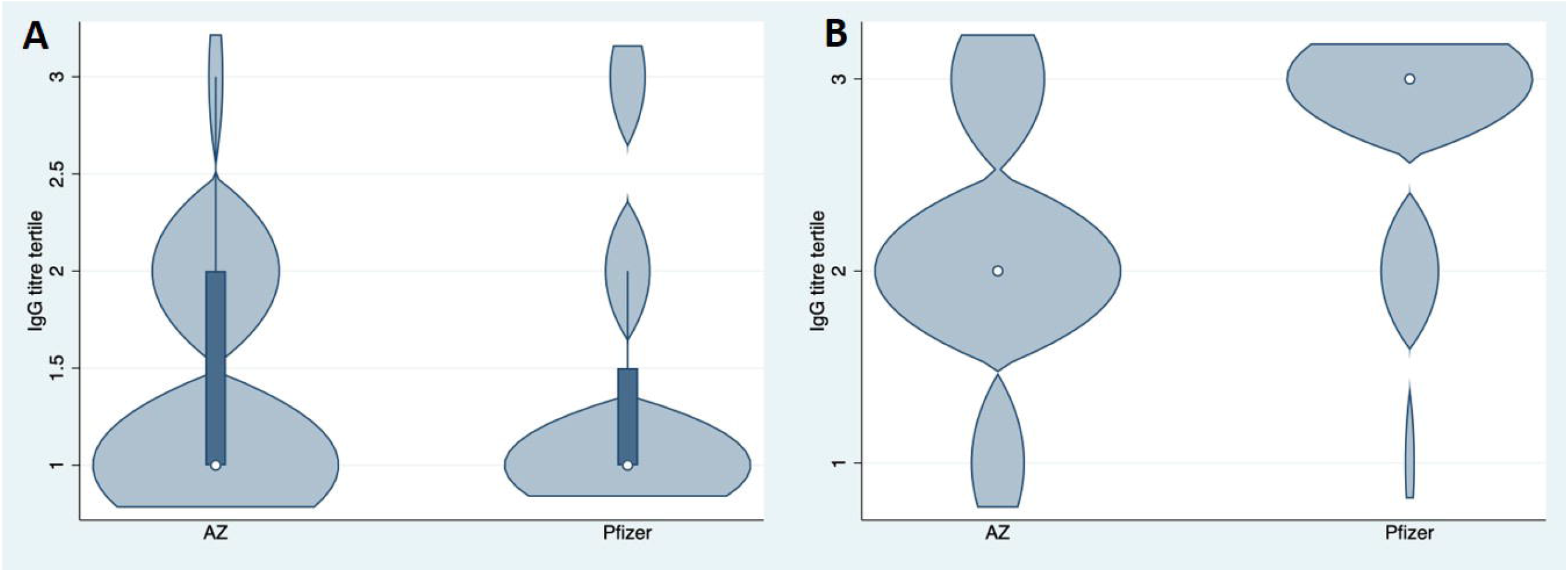
Quantitative response to vaccination in (a) those who had received anti-CD20 monoclonal antibodies, and (b) other DMT (fingolimod excluded)

## Discussion

The COVID-19 pandemic may transition to endemic and is likely to continue for many years to come, but the rapid development of vaccines offers hope for a reduction in illness severity and viral transmission, with consequent lifting of restrictions. This study answers some of the questions that have arisen around vaccine efficacy in pwMS taking immunomodulatory drugs. With a cohort of almost 500 patients, we have demonstrated that both DMT types and vaccine type affect the humoral immune response to COVID-19 vaccination. However, certain DMTs, including some with high MS treatment efficacy, appear to have no effect on the response to COVID-19 vaccination. The attenuated vaccine response observed in people on certain DMTs prompts the need to provide advice for ongoing infection prevention as the population spread of COVID-19 continues. More data is needed on whether booster vaccination improves the chance of seroconversion in people with an attenuated humoral immune response, as well as the relative importance and meaning of both humoral and T cell immunity in terms of symptomatic and severe infection.

These results align with a previous, smaller study [9], showing attenuated response to the BNT162b2 mRNA (Pfizer-BioNTech) vaccine in pwMS receiving ocrelizumab and fingolimod. The number of patients in our cohort allows us to interpret results with more certainty, examine the impact of dosing time, and importantly understand the impact of vaccine type. Anti-CD20 monoclonal antibodies were the DMTs most strongly associated with lack of seroconversion following COVID-19 vaccination. Their high efficacy and relatively favourable safety profile has offered a welcome addition to the MS DMT landscape; however, once ocrelizumab is commenced, rapid and profound B-cell depletion occurs, with few people repopulating B-cells prior to their 6 month interval infusion [10] [11]. These drugs have been shown to be associated with attenuated humoral response to other vaccinations [2], both in terms of recall and neo antigens. This presents an opportunity to test people on certain DMTs for evidence of secondary antibody deficiency both by routinely measuring Ig classes, but also by measuring functional immunity to vaccines for which normative reference ranges exist e.g. pneumococcus and haemophilus.[12]

We also found that fingolimod, a sphingosine-1-phospohate (S1P) receptor modulator, is associated with substantially attenuated humoral response to COVID-19 vaccination. This does not reflect the moderately attenuated vaccine responses seen in clinical trials [13], but has been previously demonstrated in smaller real-world studies of COVID-19 vaccine response [9]. Given the substantial MS rebound/reactivation associated with fingolimod cessation [14], medication withdrawal to facilitate successful vaccination is not feasible or ethical without a managed treatment switch to an alternative DMT. This strategy carries additional complexities including the potential for carryover DMT-associated risk e.g. lymphopenia, risk of breakthrough MS disease activity during a switch and uncertainties over how to manage a switch back if appropriate. It seems likely that the attenuated vaccine response seen with fingolimod is a class effect that will be seen in other S1P products including siponimod and ponesimod; however the different half-lives of these different medications may impact on this, meaning that drug-specific studies are needed.

Our data offers the opportunity to tailor advice for pwMS who are at risk of attenuated humoral response to vaccination. Our study suggested that delaying commencement of fingolimod or anti-CD20 DMT should be considered in new starters, to allow time for vaccination. Our data did not demonstrate a relationship between delaying established anti-CD20 infusions and improving seroconversion but this analysis is likely to have been underpowered. We did demonstrate a modest (but significant) impact of delaying established anti-CD20 infusions on COVID-19 antibody titre, raising questions over whether substantially extended dosing intervals to allow B-cell recovery might enhance vaccine response. The proportion of individuals who seroconverted increased between vaccine 1 and 2, in keeping with studies in the healthy control population [15], supports the use of booster vaccination, particularly in those who may have had an attenuated response to initial vaccination. However, further study is required to determine whether booster vaccination induces humoral immunity in all those who failed to mount a humoral response to the first course. It is not yet routine to test anti-SARS-CoV2 antibodies in clinical practice, but knowledge of seronegative status in pwMS may allow more vigilant infection control precautions e.g. continuing to socially distance and ensuring that household contacts are vaccinated, and may also allow individualised treatment of COVID-19 should they become infected e.g. monoclonal antibodies to SARS-CoV2 have been shown to benefit those who are seronegative.[16]

Our overall finding of a differential humoral response according to vaccine type replicates data in healthy control cohorts [17]. However, we did not replicate this finding in people on anti-CD20 DMT, providing no rationale for recommending one vaccine type over another in those at particular risk of attenuated response. This is deserving of further study given potential limitations of power to address this question. Current evidence suggests that heterologous vaccine regimens may elicit stronger antibody and T-cell responses [18,19]. The effect of age on vaccine response in this cohort is in line with data from the general population [15], and may be entirely due to age-related immunosenescence [17], or there may be an additional impact from collider bias related to age, disability progression and DMT choice.

This study made use of dried blood spot sampling, reducing the need for potentially vulnerable people to attend healthcare facilities during the pandemic as well as reduced costs for phlebotomist time and equipment. Dried blood spot sampling has been used since the 1960s for neonatal screening for inborn errors of metabolism [20], and has demonstrated utility across a number of medical uses [21]. Their use as a tool for serological screening of IgG levels is particularly well established [22], and the utility has been highlighted during the COVID-19 pandemic [23]. Whilst extensive work was undertaken to develop and validate the assays used in this study, the lack of gold standard for RBD assay development was a potential limitation; since a true negative result was defined based on historical samples. In order to increase the power for this study, samples were analysed in two laboratories and data pooled. The assay used in the UHW laboratory was subject to significant ceiling effects, with around a third of untreated patients having samples with an IgG titre at or above ceiling. While subgroup analysis demonstrated similar seroconversion between the two assays, and the use of tertiles avoided ceiling effects and pooling two non-linear scales, this represents a potential limitation of the study.

A further caveat is that while all DMT exposures were part of monotherapy regimens, some people with MS had been exposed sequentially to several therapies. Given the large number of DMTs, we were not powered to explore the effect of sequential therapies, so it cannot be wholly excluded.

Pre-vaccine samples were only available for 58/473 participants in this study, of whom 10% had evidence of prior infection. It is likely that at least a similar proportion of all participants already had anti-SARS-CoV-2 antibodies at baseline. There is evidence of an association between the presence of antibodies to SARS-CoV-2 at the time of vaccination and a greater subsequent immune response to the COVID-19 vaccine.[24] Given the lack of association between prior reported COVID-19 symptoms and baseline seropositivity in this cohort, we did not feel confident to use this as a marker of prior infection to explore enhanced vaccine response.

SARS-CoV-2 is a strongly immunogenic virus that can induce antigen-specific antibody production in the majority of infected patients [25]. However, the immune correlates of protection following COVID-19 vaccines have not yet been fully established. Antibody responses to COVID-19 vaccination appear to be protective[26]. Specifically, anti-SARS-Cov-2-RBD IgG levels appear to correlate with virus neutralization titres [27-29]. Durable antibody responses following COVID-19 infection have been shown to be associated with more rapid recovery from infection [30]. In animal models, neutralizing antibody (NAb) levels following vaccination using the spike antigen correlate with protection against SARS-CoV-2 [31-32], and antibody dependent functional immunity (e.g. antibody-dependent monocyte phagocytosis) is enhanced by a COVID-19 vaccine [33]. Although NAbs are frequently considered a key component of the immune response after viral infection, anti-viral T cell mediated immunity is also central to viral clearance [34]. SARS-CoV-2-specific CD4^+^ and CD8^+^ T cells have been associated with milder disease in acute and convalescent individuals [35]. Whilst we did not measure T-cell responses to the COVID-19 vaccine in this cohort, the strength of RBD-specific CD4+ T cell responses (but not CD8+ T cell) to COVID-19 infection or vaccination have been shown to correlate positively with RBD-binding IgG antibody titres[27,36]. Ongoing/future work on durability and levels required to protect from severe disease is needed in this and other cohorts.

In conclusion, this study demonstrates the impact of DMT type, vaccine type and age on vaccine response. It provides high quality evidence to support advice for people with MS, and indicates routes for future study, including the need for clinical trials to guide advice around balancing risks and potential benefits of suspending or delaying treatment.

## Data Availability

Consideration of data sharing will be made on application to the corresponding author.

## Acknowledgements

The authors would like to acknowledge the help of Swee Vickie Nixon, Ray Wynford-Thomas, Marija Cauchi, Catherine McConnell and Cynthia Butcher in the running of the study.

## Author contributions

ECT, RD, SJ, NPR, GG, KS, DB, MW, NV and SM were involved in conception, design and interpretation of the study. NV, VA, ANA, RC, NE, KG, LG, KEH, AH, GI, MJ, ASK, SJM, SNS, JS and JS were all involved in acquisition and analysis of data. ECT, RD, SJ and NPR contributed to drafting a significant portion of the manuscript or figures. Statistical analysis was performed by JB.

## Potential Conflicts of Interest

ECT has received speaker fees, consultancy fees or travel expenses to attend educational meetings from Merck, Biogen, Roche and Novartis.

NV, ANA, RC, MJ, JB, SNS, KG, BD, SL, AH, LG, GI, VA, KB, JS and MU have no conflicts of interest.

AK has trademarked GloBody and filed patents for potential commercial development related to the GloBody technology

KS has received speaker honoraria from Novartis, Biogen, Teva, Merck KG Aa, Sanofi-Genzyme, and Roche; sat on advisory boards for Novartis, Merck KG Aa, and Roche. He has received grant support from Biogen and Merck.

GG has received speaker honoraria from AbbVie, Actelion, Biogen, Celgene, Sanofi-Genzyme, Genentech, Merck-Serono, Novartis, Roche and Teva; sat on advisory boards for AbbVie, Actelion, Biogen, Celgene, Sanofi-Genzyme, Genentech, GlaxoSmithKline, Merck-Serono, Novartis, Roche and Teva; and received research support from Sanofi-Genzyme, Takeda, and Merck

SJ has received support for conferences, speaker, advisory boards, trials, Data and Safety Monitoring Boards, and projects with CSL Behring, Takeda, Swedish Orphan Biovitrum, Biotest, Binding Site, Grifols, BPL, Octapharma, LFB, Pharming, GSK, Weatherden, Zarodex, Sanofi, and UCB Pharma. None of these conflicts relate to the current work.

RD has received speaker honoraria from Biogen Idec, Teva, Merck, Neurology Academy and Sanofi Genzyme; sat on advisory boards for Roche, Merck and Biogen; and received research support from Biogen, Merck and Celgene.

**Supplementary table 1:**
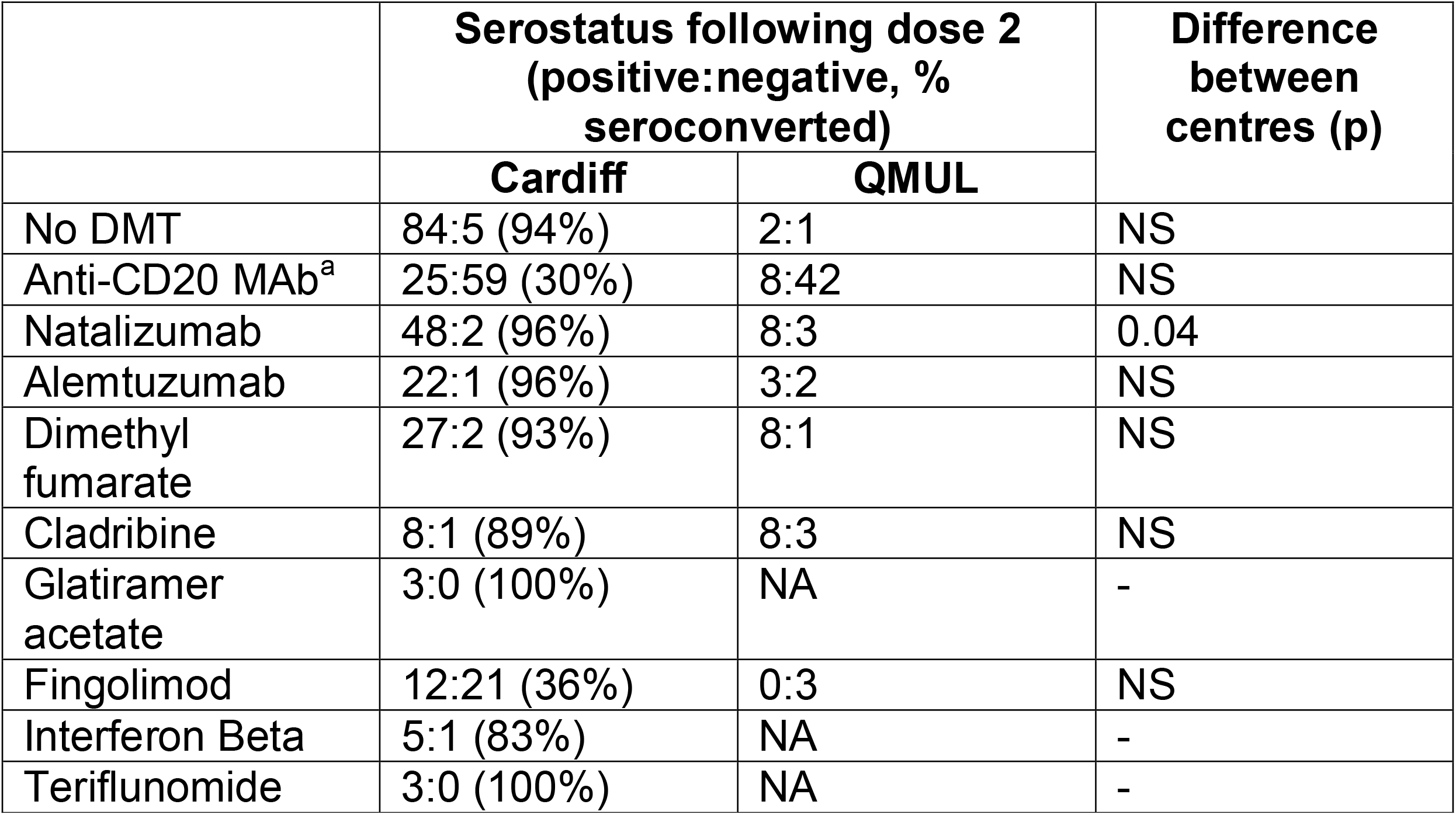
Samples grouped by site of analysis

